# Genetic overlap between Parkinson disease and inflammatory bowel disease

**DOI:** 10.1101/2022.01.13.22269221

**Authors:** Xiaoying Kang, Alexander Ploner, Yunzhang Wang, Jonas F. Ludvigsson, Dylan M. Williams, Nancy L. Pedersen, Karin Wirdefeldt

## Abstract

**Importance:** Parkinson disease (PD) and inflammatory bowel disease (IBD) have been associated, implying shared pathophysiology. Characterizing genetic pleiotropy between the two conditions aids the exploration of common etiology.

**Objective:** To estimate the genetic correlation between PD and IBD and to identify specific loci influencing both conditions.

**Design:** Genetic study with applications of high definition likelihood and conditional false discovery rate (FDR) framework.

**Setting:** The study was based on summary statistics of genome-wide association studies (GWAS).

**Participants:** The PD GWAS comprised 37,688 cases and 981,372 controls, and the IBD GWAS included 25,042 cases and 34,915 controls. Participants were of mixed ethnicity.

**Exposures:** None.

**Main Outcomes and Measures:** The main outcomes were a set of single nucleotide polymorphisms (SNPs) identified by conditional FDR analysis as jointly associated with PD and IBD.

**Results:** Weak but statistically significant genetic correlations were detected for PD with both Crohn’s disease (CD) and ulcerative colitis (UC), the two main subtypes of IBD. A total of 1333 SNPs in 28 genomic loci and 1915 SNPs in 22 loci were jointly associated with PD-CD and PD-UC, respectively, at conjunctional FDR under 0.01. The pleiotropic loci appeared distinctive for PD-CD and PD-UC, are mostly novel and comprise loci with either same or opposing genetic effects on the two phenotypes. Positional and eQTL mapping prioritized 316 PD-CD and 303 PD-UC genes, among which only <10% are differentially expressed in both colon and substantia nigra. The KEGG pathways enriched by all prioritized genes were highly concordant between PD-CD and PD-UC, with the majority being related to immune and/or autoimmune dysfunction.

**Conclusions and Relevance:** Overall, we found robust evidence for a genetic link between PD and each subtype of IBD. The identified genetic overlap is complex at the locus and gene levels, indicating the presence of both common etiology and antagonistic pleiotropy. At the functional level, our results highlighted a central role of host immunity and/or autoimmunity in the PD-IBD relationship.

## Introduction

Parkinson disease (PD) is a neurodegenerative movement disorder with no curative or disease-modifying therapies, suggesting an urgent need for better understanding of disease pathophysiology to foster drug discovery. The gut-brain axis has been hypothesized to play a role in PD pathogenesis, stimulating a growing body of work on the putative contribution of gastrointestinal dysfunction in PD initiation.^1,2^ Inflammatory bowel disease (IBD) is a chronic intestinal inflammatory condition manifested by long-lasting diarrhea, abdominal pain and bloody stool.^3^ Recently, a meta-analysis of nine observational studies comprising over 12 million patients demonstrated an interesting bidirectional relationship between PD and IBD, as well as a protective effect of anti-inflammatory medications on PD among IBD patients.^4^ The observed IBD-PD correlation was further strengthened by subsequent epidemiological findings, which collectively indicated a biological link between the two seemingly unrelated conditions.^5,6^ The onset of PD-like neuropathology and motor function impairment in an animal model for IBD also supported the involvement of intestinal inflammation in PD initiation and progression.^7^

How the inflamed gut and its underlying mechanisms are intrinsically connected to PD remains elusive. A sophisticated interplay between mucosal immunity and intestinal microbiota, two key drivers in IBD, has been shown to be relevant.^8-10^ Mechanistically, the inflammation-associated disruption of the intestinal epithelial barrier facilitates the translocation of microbial products from the intestinal lumen into the peripheral circulation, inducing gut dysbiosis and systemic inflammation.^11^ These events next trigger upregulation of α-synuclein (the PD pathogenic protein) expression and its abnormal aggregation in enteric neurons, as well as neuroinflammation, which promotes neurodegeneration in the brain.^12,13^ Discovery of genetic overlap between PD and IBD has shed new light into the molecular underpinning shared by the two diseases. For instance, polymorphisms of the leucine-rich repeat kinase 2 (*LRRK2*) gene have been robustly associated with susceptibility of both PD and Crohn’s disease (CD), a subtype of IBD, corroborating the crucial role of the immune system in the two conditions.^14,15^ Owing to the methodological advancement in cross-phenotype pleiotropic analyses, additional common genetic determinants of PD and IBD have been identified.^16^ Hence, to further test the PD-IBD connection genetically and to uncover potential novel mechanistic explanations for this relationship, we evaluated the genetic correlation between the two diseases and characterized their cross-trait pleiotropic profiles using summary statistics from updated genome-wide association studies (GWAS) on PD and IBD.

## Methods

### Source data

Summary statistics from two GWAS were analyzed in the present work. Genetic associations with PD were derived from a meta-analysis of 16 case-control samples from the International Parkinson’s Disease Genomics Consortium (IPDGC) and 23andMe, Inc; comprising a total of 37,688 patients with clinically ascertained (65%) or self-reported (35%) PD and 981,372 controls. Sample characteristics and the study protocol are described elsewhere.^17^

The IBD GWAS summary statistics were based on a total of 59,957 participants, combining a UK sample of 25,305 European individuals (N_case/control_ = 12,160/13,145) and the International Inflammatory Bowel Disease Genetics Consortium sample (N_case/control_ = 12,882/21,770).^18^ All IBD patients were clinically ascertained, and subtype information on CD or ulcerative colitis (UC) was available. Details about study participants and protocol can be found in the original publication.^18^ Given the pathophysiological and genetic differences between IBD subtypes^19-21^, we accessed the genetic associations with both CD (combined N_case/control_ = 12,194/28,072) and UC (combined N_case/control_ = 12,366/33,609), and analyzed the two subtypes separately throughout the study.

### Genetic correlation analysis

The analytical process is displayed as a flowchart in Figure 1. Genetic correlations (rg) between PD and each IBD subtype were estimated via high-definition likelihood (HDL), a full likelihood-based extension of the conventional LD score regression method that improves rg estimation precision by integrating more information on the linkage disequilibrium (LD) structure.^22,23^

**Figure 1.**
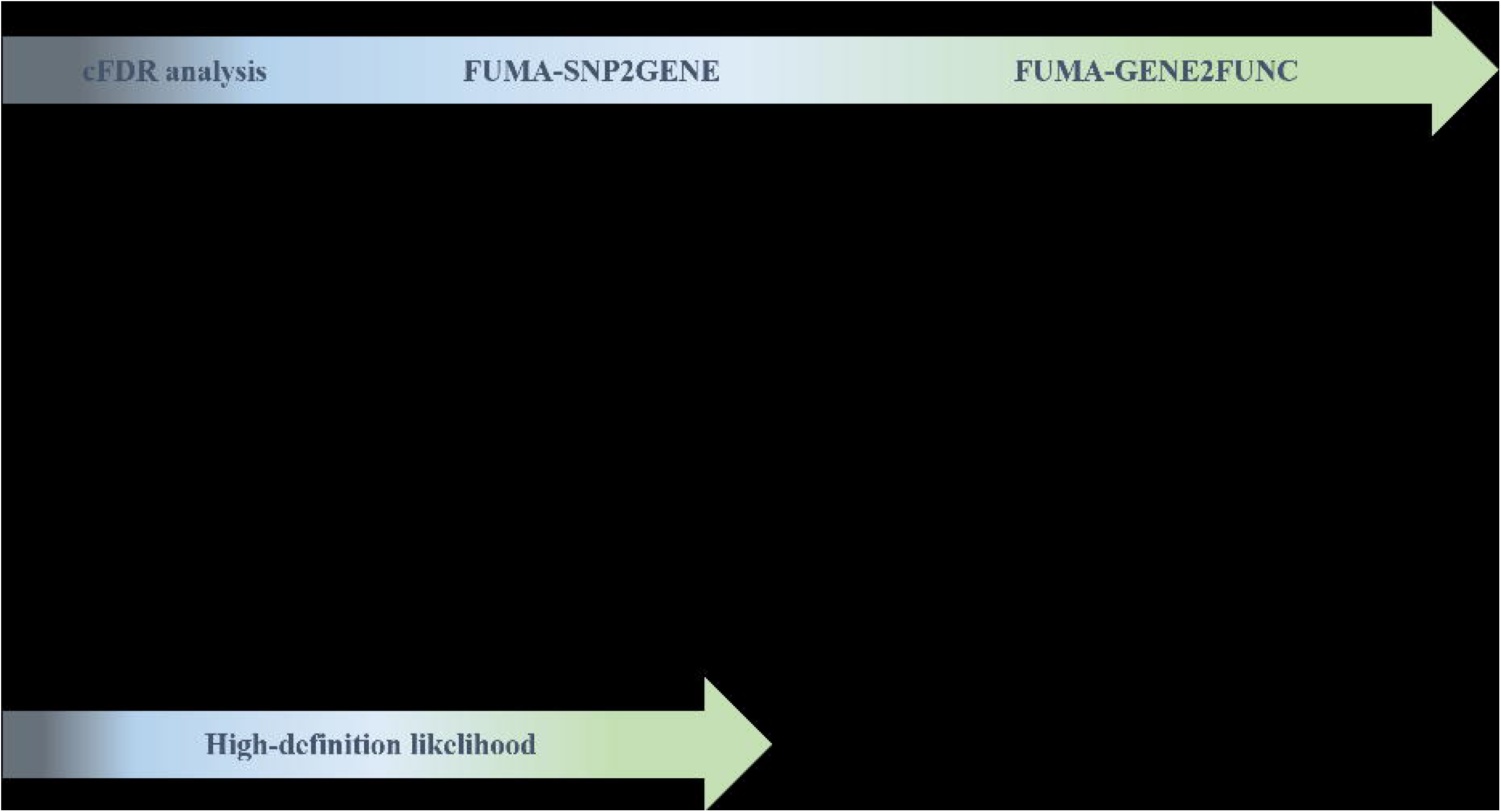
Schematic diagram of the analytical procedures implemented in the study. *Abbreviations*: cFDR, conditional false discovery rate; PD, Parkinson disease; IBD, inflammatory bowel disease; SNP, single nucleotide polymorphism.

### Conditional false discovery rate analysis

Genetic overlap between PD and each IBD subtype was explored using the conditional false discovery rate (cFDR) framework proposed by Andreassen et al.^24^ First, we visualized the extent of shared genetics affecting both traits as conditional or stratified quantile-quantile (Q-Q) plots: for any single phenotype, a conventional Q-Q plot shows the quantiles of the - log10-transformed *p*-values from the corresponding GWAS (on the vertical axis) against the quantiles from an equally transformed uniform distribution corresponding to a global null distribution (on the horizontal axis); an upward deflection of the resulting curve from the line of identity reflects an enrichment of smaller than expected *p*-values, corresponding to a deviation from the null and the presence of a biological signal. For pairs of phenotypes, conditional Q-Q plots show multiple such quantile curves for the primary or target phenotype, where each curve corresponds to a subset of variants where the *p*-values for the secondary or conditioning phenotype are selected by increasingly stringent thresholds: in the complete absence of pleiotropy, these curves should coincide, whereas in the presence of pleiotropy, we expect to see increasing upward deflection from the global null, and consequently increasing enrichment of variants associated with the primary phenotype among the subsets defined by the secondary phenotype.^24-26^

Next, we identified pleiotropic SNPs for PD-CD and PD-UC, respectively, by calculating the conjunctional false discovery rate (conjFDR) value for each SNP included in both PD and IBD datasets. The conjFDR is based on the cFDR and can be interpreted as a conservative estimate of the false discovery rate for a given SNP being jointly associated with both phenotypes under investigation.^24^ We defined pleiotropic SNPs as those with conjFDR below a threshold of 0.01.

To assure approximate independence of the variants/*p*-values involved, we implemented LD-based random pruning for both conditional Q-Q plots and calculation of the conjFDR (the latter based on 100 independent iterations).^24,25^ Further details about the concept and the recommended analytical protocol for cFDR analysis are reviewed by Smeland et al.^25^

### Functional mapping and annotation of pleiotropic variants

Characterization and functional annotation of the pleiotropic SNPs identified from cFDR were performed via the FUMA web-based platform under the default setting if not otherwise specified.^27^ Using the FUMA-SNP2GENE function, we first annotated all pleiotropic SNPs via the built-in ANNOVAR tool and identified their corresponding genomic loci based on the LD pre-computed from the 1000 Genome reference panel.^28,29^ The direction of genetic effects of a pleiotropic locus on PD and IBD subtype was evaluated based on the proportion of concordant SNPs, defined as the variants with the product of two association coefficients β_PD_ β_IBD_ > 0 among the total SNPs within the locus. We regarded a locus containing ≤10%, 10-90%, and ≥90% of concordant SNPs as being of antagonistic, ambiguous and concordant pleiotropy, respectively. The distribution of jointly associated SNPs and loci in different directions of pleiotropy were then visualized as Manhattan plots. To identify novel pleiotropic loci that have never been previously associated with both PD and CD, or PD and UC, we searched the PD GWAS Locus Browser (https://pdgenetics.shinyapps.io/GWASBrowser/)^30^ and the GWAS Catalog (https://www.ebi.ac.uk/gwas/) for all shared loci informed by our cFDR analysis, considering evidence from both GWAS and cross-phenotype analysis.

Next, we proceeded with gene prioritization via positional and expression quantitative trait loci (eQTL) mapping. We selected the GTEx v8 database for eQTL mapping, and restricted the tissue types to colon (including sigmoid and transverse) and substantia nigra, which are most relevant to IBD and PD pathophysiology.^31^ All pleiotropic SNPs were gene-mapped using the FUMA-SNP2GENE function without any filtering per functional annotations (i.e. functional consequence or Combined Annotation Dependent Depletion, CADD score). The genes prioritized via positional and eQTL mapping were then taken forward to the FUMA-GENE2FUNC function to infer putative biological pathways by over-representation analysis. Significantly enriched gene sets or pathways were determined per FUMA-GENE2FUNC default parameters adjusting for multiple testing.

### Standard Protocol Approvals, Registrations, and Patient Consents

For each GWAS study included in the present work, written informed consent was received from all participants and ethical approval was obtained from relevant ethical review boards. No additional ethical approval was required for our study because the analyses were based on summary statistics – i.e. without accessing individual-level genetic data.

### Data availability

Full summary statistics of GWAS on IBD are publicly available at GWAS Catalog: genetic associations with CD can be downloaded at https://www.ebi.ac.uk/gwas/studies/GCST004132 and for UC at https://www.ebi.ac.uk/gwas/studies/GCST004133. Summary statistics of PD GWAS can be obtained via research project applications to 23andMe and the IPDGC. For 23andMe, the full GWAS summary statistics for the discovery data set will be made available through 23andMe to qualified researchers under an agreement with 23andMe that protects the privacy of the 23andMe participants. Please visit research.23andme.com/collaborate/ for more information and to apply to access the data.

### Code availability

HDL analysis was performed via the software provided by Ning Z et al. at https://github.com/zhenin/HDL/. The cFDR analysis was performed with R version 4.0.3 using the “cfdr.pleio” package available at https://github.com/alexploner/cfdr.pleio.

## Results

Weak but statistically significant genetic correlations of PD with both CD (*r*_*g*_ = 0.06, s.e. = 0.02; *P* = 0.01) and UC (*r*_*g*_ = 0.06, s.e. = 0.03; *P* = 0.03) were detected by HDL (Table 1). In accordance, the successive leftward shift of curves seen in the conditional QQ plots for both trait pairs in both directions corroborated the presence of genetic overlap between PD and each IBD subtype (Figure S1). These deflections can be understood as increases in the excess of non-null SNPs for the principal phenotype when sequentially selecting sets of SNPs with stronger evidence of conditional associations. Interestingly, we noticed that the curves separated more prominently when more relaxed *p* levels were conditioned on. Once the conditional *p* cutoff reached 1×10^−5^ (represented by the green curve in Figure S1), there was not much gain in association enrichment by requiring stronger evidence on conditional associations. Such trends might imply that the pleiotropy underlying PD and IBD is mainly attributable to SNPs with mild to moderate levels of evidence on phenotype associations but not to those with top signals for each trait.

**Table 1.** Genetic correlations between Parkinson disease and each subtype of inflammatory bowel disease.

| Phenotypes | $r_g$ | *Standard error* | $p$ |
| --- | --- | --- | --- |
| PD and CD | 0.06 | 0.02 | 0.01 |
| PD and UC | 0.06 | 0.03 | 0.03 |
| *Abbreviations:* PD, Parkinson disease; CD, Crohn disease; UC, ulcerative colitis. |  |  |  |

Using cFDR analysis, we identified 1,333 and 1,915 SNPs at conjFDR below 0.01 for PD-CD and PD-UC, respectively, of which 413 were common to both trait pairs (Table 2). Overall, these jointly associated SNPs were mostly concordant with same genetic effects on the two phenotypes (72.2% for PD-CD and 75.8% PD-UC), located in non-exonic regions (96.8% for PD-CD and 97.8% for PD-UC), and less likely to be deleterious due to a CADD score under 12.37 (96.4% for PD-CD and 96.1% for PD-UC).^32^ The pleiotropic SNPs were then mapped to 28 independent loci for PD-CD and 22 for PD-UC, with 11 pairwise overlapping loci (Table S1-2). Amongst the 39 distinct loci in total, 30 had never been previously reported to affect both PD and IBD or either of its subtypes, and were therefore considered as novel shared loci (Table S5). As displayed in the Manhattan plots, the locus-level pleiotropic patterns for PD-CD and PD-UC exhibited similarities in that the jointly associated loci were distributed widely across the entire genome, with at least half affecting the PD/IBD risk in the same direction; additionally, the ambiguous locus *ARIH2* for PD-CD locates in the same region as the ambiguous PD-UC locus at *IP6K2* (Figure 2, Table S5). The two trait pairs however differed in their strongest association, which was located in the *SLC2A13* locus on chromosome 12 (shared with *LRRK2*) for PD-CD, but in the *HLA-DRA* locus on chromosome 6 for PD-UC. Interestingly, three of the 11 pairwise common loci had different pleiotropic directions: *IL1R2* on chromosome 2 and *HIST1H2BO* (shared with PD-UC locus *ZNF165*) on chromosome 6 were antagonistic for PD-CD but concordant for PD-UC, while *TRIM10* (shared with PD-UC locus *HLA-W*) on chromosome 6 was concordant for PD-CD but antagonistic for PD-UC. The discrepancy of pleiotropic direction for common loci mirrors the earlier finding that CD and UC may be genetically distinctive.^20^

**Table 2.** Genome-wide pleiotropy for Parkinson disease and inflammatory bowel disease.

|  | PD and Crohn disease | PD and ulcerative colitis |
| --- | --- | --- |
| *Pleiotropic SNPs at $\text{conjFDR} < 0.01$* |  |  |
| Total, n (%) | 1,333 (100%) | 1,915 (100%) |
| Concordant SNPs, n (%) | 962 (72.2%) | 1,452 (75.8%) |
| Exonic SNPs, n (%) | 42 (3.2%) | 42 (2.2%) |
| SNPs with CADD > 12.37, n (%) | 48 (3.6%) | 74 (3.9%) |
| *Independent loci identified from the pleiotropic SNPs, by direction of pleiotropy<sup>1</sup>* |  |  |
| Total, n (%) | 28 (100%) | 22 (100%) |
| Concordant, n (%) | 14 (50.0%) | 13 (59.1%) |
| Antagonistic, n (%) | 13 (46.4%) | 8 (36.4%) |
| Ambiguous, n (%) | 1 (3.6%) | 1 (4.5%) |
| *Prioritized genes, by mapping method* |  |  |
| Total | 316 | 303 |
| eQTL, colon only<sup>2</sup> | 112 | 100 |
| eQTL, substantia nigra only | 2 | 1 |
| eQTL, colon & substantia nigra | 21 | 25 |
| *Over-represented gene sets or pathways with adjusted $p < 0.05$<sup>3</sup>* |  |  |
| KEGG pathways | 12 | 16 |
| GO term: biological process | 62 | 78 |
| GO term: cellular component | 41 | 46 |
| GO term: molecular function | 6 | 12 |

<sup>1</sup>Direction of pleiotropy for a locus is defined by the proportion of concordant SNPs divided by the total number of SNPs within the locus: concordant if >90%, antagonistic if <10%, and ambiguous if 10-90%.
<sup>2</sup>Includes sigmoid and transverse
<sup>3</sup>Based on genes identified by both positional and eQTL mapping via FUMA-GENE2FUNC.
*Abbreviations:* PD, Parkinson disease; SNP, single nucleotide polymorphism; conjFDR, conjunctural false discovery rate; CADD, combined annotation dependent depletion; eQTL, expression quantitative trait loci; GO, gene ontology; KEGG, kyoto encyclopedia of genes and genomes.

**Figure 2.**
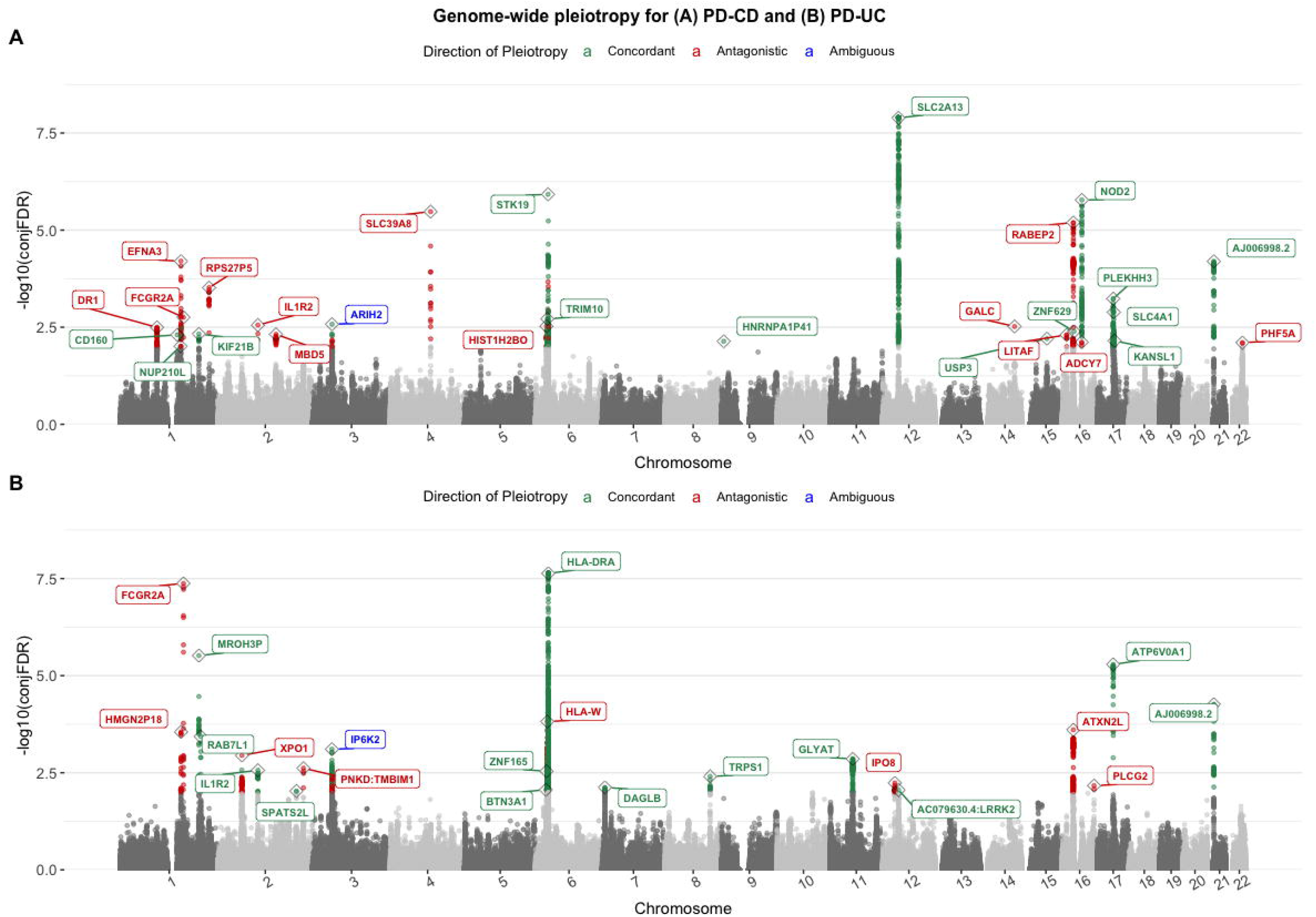
Manhattan plots for jointly associated variants and loci in different directions of pleiotropy for Parkinson disease and Crohn’s disease (A) or ulcerative colitis (B). The y-axis denotes the -log10-transformed conjunctional false discovery rate (conjFDR) values for each single nucleotide polymorphism (SNP). The jointly associated SNPs with conjFDR < 0.01 were highlighted in green if being concordant (with same genetic effects on the two phenotypes), or red if being antagonistic (with opposite genetic effects on the two phenotypes). The jointly associated loci were labelled with the corresponding gene name, and colored in green, red or blue if the direction of locus pleiotropy is concordant, antagonistic, or ambiguous, respectively. *Abbreviations*: PD, Parkinson disease; CD, Crohn’s disease; UC, ulcerative colitis; conjFDR, conjunctional false discovery rate.

Positional and eQTL mapping prioritized 316 and 303 genes for PD-CD and PD-UC, respectively, among which 197 pairwise overlapped (Table 2, S3-4). For both trait pairs, around 80% of the eQTL mapped genes are differentially expressed in colon but not substantia nigra. Following the FUMA-GENE2FUNC procedure, 12 KEGG pathways for PD-CD and 16 for PD-UC (Figure 3, Table S6), as well as a wide range of Gene Ontology (GO) terms (results not shown) were found to be over-represented by all the genes prioritized from both mapping methods and including both tissues. Comparison of KEGG results showed that 11 of the 12 PD-CD pathways, all related to host immunity and/or autoimmune diseases, were also significantly enriched in PD-UC genes; whereas the “KEGG_LYSOSOME” pathway, which comprised one of the most common PD susceptibility gene *GBA*, was identified for PD-CD but not PD-UC. Beyond the immune-dominated mechanisms, the pathways specific to PD-UC further extended to neuronal development and hematopoietic development.

**Figure 3.**
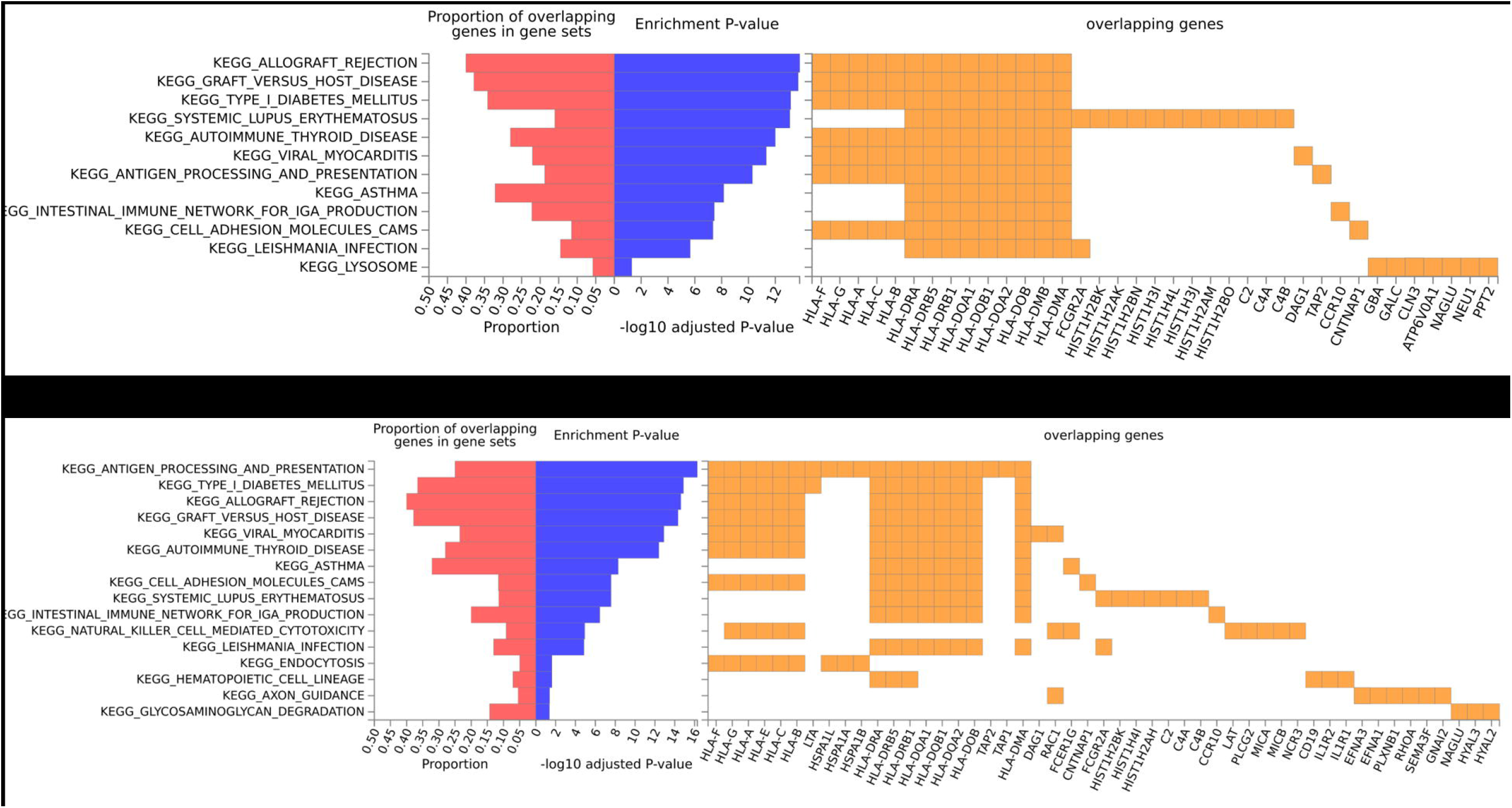
The KEGG pathways over-represented by prioritized genes for Parkinson disease and Crohn’s disease (A) or ulcerative colitis (B). The -log10 adjusted *P*-value denotes the - log10-transformed *P*-value from gene-set enrichment analysis after multiple test correction. *Abbreviations*: PD, Parkinson disease; CD, Crohn’s disease; UC, ulcerative colitis.

## Discussion

Despite a modest genetic correlation, we discovered robust evidence for a genetic link between PD and each IBD subtype, underpinned by many shared genomic loci. The identified genetic overlap is complex at the locus- and gene-levels, indicating the presence of both common etiology and antagonistic pleiotropy between PD and IBD. Nonetheless, at the functional level, the PD-IBD genetic overlap converges to a set of pathways involved in host immunity and/or autoimmunity.

The biological connection between PD and IBD has been supported by a growing body of evidence.^4,14,16^ The genetic correlation between PD and each IBD subtype however was weak in our material (albeit conventionally statistically significant), in contrast to the evidence for pleiotropic enrichment seen in the conditional Q-Q plots. Possible explanations for this unexpectedly weak genetic correlation may be found in the cFDR results. First, the PD-IBD pleiotropy was observed to be mainly due to SNPs with GWAS *p*-values above the genome-wide significance level, whereas such non-significant variants are largely under-weighted in the estimation of genetic correlation.^22^ Second, the genetic overlap between PD and each IBD subtype comprises antagonistic pleiotropy, which may cancel out the effects of shared risk loci and lead to an underestimated genetic correlation.^33^ Taken together, these two points emphasize the importance of including not only SNPs with genome-wide significance but also those with moderate association *p*-values, and considering the direction of genetic effects in future explorations of pleiotropy for complex traits.

Several previously established shared loci, such as *HLA* and *LRRK2*, are replicated by our findings. Moreover, we discovered 30 novel pleiotropic loci, including 10 known risk loci for either PD (*RAB7L1* and *ARIH2/IP6K2*) or IBD (*KIF21B/MROH3P, IL1R2, HNRNPA1P41, GLYAT, PNKD:TMBIM1, LITAF, ADCY7* and *PLCG2*).^17,18,34-40^ Amongst these, *MROH3P* is an IBD risk locus that was initially nominated as a shared locus for PD-CD but not PD-UC.^16^ Here, we confirmed its concordant pleiotropy for PD-CD and extended it further to PD-UC. The association of *MROH3P* with colonic expression of *C1orf106*, an IBD susceptibility gene encoding a protein that is critical for epithelial homeostasis, suggests a role of intestinal barrier dysfunction in PD and IBD pathogenesis.^41^ For another IBD risk locus *IL1R2*, which harbors the encoding gene for interleukin-1 receptor 2, we found conflicting pleiotropy for PD-CD (antagonistic) versus PD-UC (concordant). This is not intuitive because the immune regulatory role of interleukin-1 receptor 2 in IBD has been well documented.^42^ Interestingly, emerging data also associated *IL1R2* polymorphisms with PD risk.^43-45^ Although our eQTL mapping results do not support any functional impact of *IL1R2* variation on gene expression in colon or substantia nigra, future research is warranted. The PD risk locus at *ARIH2/IP6K2* is also noteworthy for its association with the expression of candidate PD gene *WDR6* and 4 other genes (*NCKIPSD, GMPPB, PRKAR2A*, and *AMT*) in both colon and substantia nigra in our data.^46^ Intriguingly, the pleiotropic direction of *ARIH2/IP6K2* locus for PD-CD and PD-UC remained ambiguous in our data and needs subsequent studies to clarify. Lastly, the 20 pleiotropic loci with no documented association with any studied traits contained 4 eQTLs for expression of 6 genes in both colon and substantia nigra; however, none of them were over-represented in any KEGG gene-sets.

In contrast to the complexity of locus-level pleiotropy, the biological pathways shared by PD and IBD are dominated by immune-related mechanisms. As displayed in Figure 3, the 12 PD-CD and 16 PD-UC KEGG pathways overlapped in 11 gene-sets, where the enriched genes are predominantly HLA members that are responsible for antigen presentation and immune regulation.^47^ Notably, four autoimmune phenotypes – type 1 diabetes, systemic lupus erythematosus, autoimmune thyroid disease and asthma – were commonly identified for both PD-CD and PD-UC, in line with Witoelar et al’s earlier report.^16^ Another 5 terms were implicated in dysregulation of immune response to external stimuli, such as transplanted grafts (*allograft rejection* and *graft versus host disease*) and pathogens (*viral myocarditis, intestinal immune network for IgA production*, and *Leishmania infection*), suggesting a pivotal role of host defense mechanisms in the PD-IBD shared biology.

Strengths of our study are the implementation of the powerful contemporary methods and the utilization of the most updated GWAS data. The reproducibility of our findings is enhanced by the choice of a conservative conjFDR threshold of 0.01. To our knowledge, we are the first to make the distinction between concordant and antagonistic pleiotropy in research on the PD-IBD connection, which provides crucial information for translational clinical and pharmaceutical innovation. The present work also has limitations. First, functional validation of detected genetic elements is beyond our scope, restricting causal interpretation of our findings. Nevertheless, we performed multi-hierarchical bioinformatic analyses to facilitate biological inference. Second, based on the data available to us, we were not able to clarify the pleiotropic direction for all identified genetic overlap between PD and IBD. However, the pathway analysis indicated that the immune system remains the most promising common target for the two diseases, regardless of the sophisticated pleiotropic pattern at locus or gene levels.

In conclusion, our genetic evidence supports the notion that PD and IBD are biologically connected phenotypes and indicate the immune system as a promising target for therapeutic development for both PD and IBD.

## Data Availability

All data analyzed in the present study are publicly available summary statistics of multiple GWAS data. Full summary statistics of GWAS on IBD are publicly available at GWAS Catalog: genetic associations with CD can be downloaded at https://www.ebi.ac.uk/gwas/studies/GCST004132 and for UC at https://www.ebi.ac.uk/gwas/studies/GCST004133. Summary statistics of PD GWAS can be obtained via research project applications to 23andMe and the IPDGC. For 23andMe, the full GWAS summary statistics for the discovery data set will be made available through 23andMe to qualified researchers under an agreement with 23andMe that protects the privacy of the 23andMe participants. Please visit research.23andme.com/collaborate/ for more information and to apply to access the data.

## Acknowledgements

The authors thank Dr. Ida Karlsson from Karolinska Institutet for suggestions on choice and application of functional annotation tools, Dr. Zheng Ning from Karolinska Institutet for assistance on running the HDL package. The authors would also like to thank the research participants and employees of the IPDGC and 23andMe for making this work possible. Contributors to the IPDGC are listed at https://pdgenetics.org/partners.

## Author Contributions

X.K. had full access to the data analyzed in the study and take responsibility for the integrity of the data and the accuracy of data analysis.

*Concept and design*: X.K., A.P., and K.W..

*Acquisition, analysis, or interpretation of data*: X.K., A.P., Y.W., and K.W..

*Drafting of the manuscript*: X.K..

*Critical revision of the manuscript for important intellectual content*: All authors.

*Statistical analysis*: X.K., A.P..

*Obtained funding*: N.L.P. and K.W..

*Administrative, technical, or material support*: K.W..

*Supervision*: A.P., D.M.W., N.L.P., J.F.L., and K.W..

